# Exploring booster session attendance, prescription, and outcomes in adults with chronic low back pain: Secondary analysis of a randomized clinical trial

**DOI:** 10.1101/2025.01.27.25321189

**Authors:** Vanessa M. Lanier, Keith R. Lohse, Quenten L. Hooker, Jesse M. Civello, Linda R. van Dillen

## Abstract

**Introduction:** Booster sessions are a potential method for maintaining self-management behaviors and treatment effects in people with chronic low back pain (LBP). However, few studies have examined booster prescription or outcomes in people with LBP.

**Objective:** (1) Compare booster prescription for two exercise-based treatments for low back pain (LBP) in a randomized clinical trial (RCT) where the number of boosters prescribed was based on self-management program independence, (2) Determine if there are variables that predict who will require >1 booster, (3) Explore the effects of boosters on pain and function in people who required >1 booster.

**Design:** Secondary analysis of a RCT in which participants were randomized to motor skill training (MST), MST+Boosters (MST+B), strength and flexibility exercise (SFE), or SFE+B.

**Setting:** Academic research setting.

**Participants:** 76 participants with chronic LBP assigned to receive boosters.

**Interventions:** This secondary analysis focuses only on the MST+B and SFE+B groups. Both groups received 6 visits of MST or SFE and six months later received up to 3 boosters. The number of boosters was based on self-management program independence at the first booster. Those who required >1 booster were not able to independently perform their program at the first booster.

**Main Outcome Measures:** Booster attendance and prescription, pain (Numeric Pain Rating Scale), function (modified Oswestry Disability Questionnaire)

**Results:** There was not a significant difference between MST+B and SFE+B in returning for the initial booster, χ^2^(1)=1.76, p=0.185. SFE+B were more likely to require >1 booster than MST+B; *β*=2.39, *p*<0.001. No participant-specific factors we examined were statistically related to needing >1 booster.

**Conclusion:** MST+B participants were less likely to require additional boosters. No additional participant-specific factors we examined were associated with needing additional boosters. Qualitatively, attending additional booster sessions did not appear to change pain or function in the current sample.

## INTRODUCTION

Chronic low back pain (LBP) is a prevalent, costly condition and is the leading cause of disability in the world.^1^ Many rehabilitative interventions for LBP have been examined with positive effects on pain and function, including exercise, manual, and behavioral therapy.^2–5^ Unfortunately, the largest effects are typically attained at short-term follow up.^4,6,7^ Given that LBP is a condition with a course of persistent symptoms and disability for many, a goal is to prolong the positive effects of intervention s over time, and self-management programs of exercise, education, or psychological interventions are suggested following an initial intervention.^8^ Because long-term adherence to self-management programs is challenging, boosters, or additional sessions provided periodically following an initial intervention, have been suggested as a method for maintaining self-management behaviors and treatment effects in people with LBP. ^8–10^

Some studies have examined boosters for people with chronic LBP.^11–19^ However, in most studies, boosters were provided without a “no booster” comparison group, making it impossible to determine if there was a booster effect.^14–19^ The few studies that compared identical interventions with and without boosters did not identify a significant booster effect on pain and function.^11–13^ Additionally, the most appropriate number of booster sessions is not clear. In previous studies, there was substantial variation in the number of boosters ranging from 1 to 24 sessions and the same number of sessions was prescribed for all participants, regardless of status at the time of the booster.^11–19^ Since providing additional treatment is costly, it would be ideal to deliver the fewest sessions possible at a relevant time point to maintain self-management behaviors and treatment effects, which might vary per individual. If we could predict who would need additional boosters, then we could focus resources on those individuals.

This study is a secondary analysis of data from a randomized clinical trial (RCT) comparing motor skill training (MST), MST+Boosters (MST+B), strength and flexibility exercise (SFE), or SFE+B for chronic LBP.^20^ In the primary analysis, there were no statistically significant effects of boosters on pain or function (i.e., +B groups compared to other groups). ^20^ However, participants received different numbers of boosters based on their ability to perform their self-management program independently. Those who were not independent at the first booster were prescribed additional booster sessions (>1 booster up to 3 maximum; **FIGURE 1**). This design is novel in studies of boosters. The purposes of this hypothesis-generating study, therefore, are to (1) compare booster attendance and prescription in the MST+B and SFE+B groups, (2) determine if there are variables that predict who needs >1 booster, and (3) explore the effects of boosters on pain and function in people who needed >1 booster.

**FIGURE 1.**
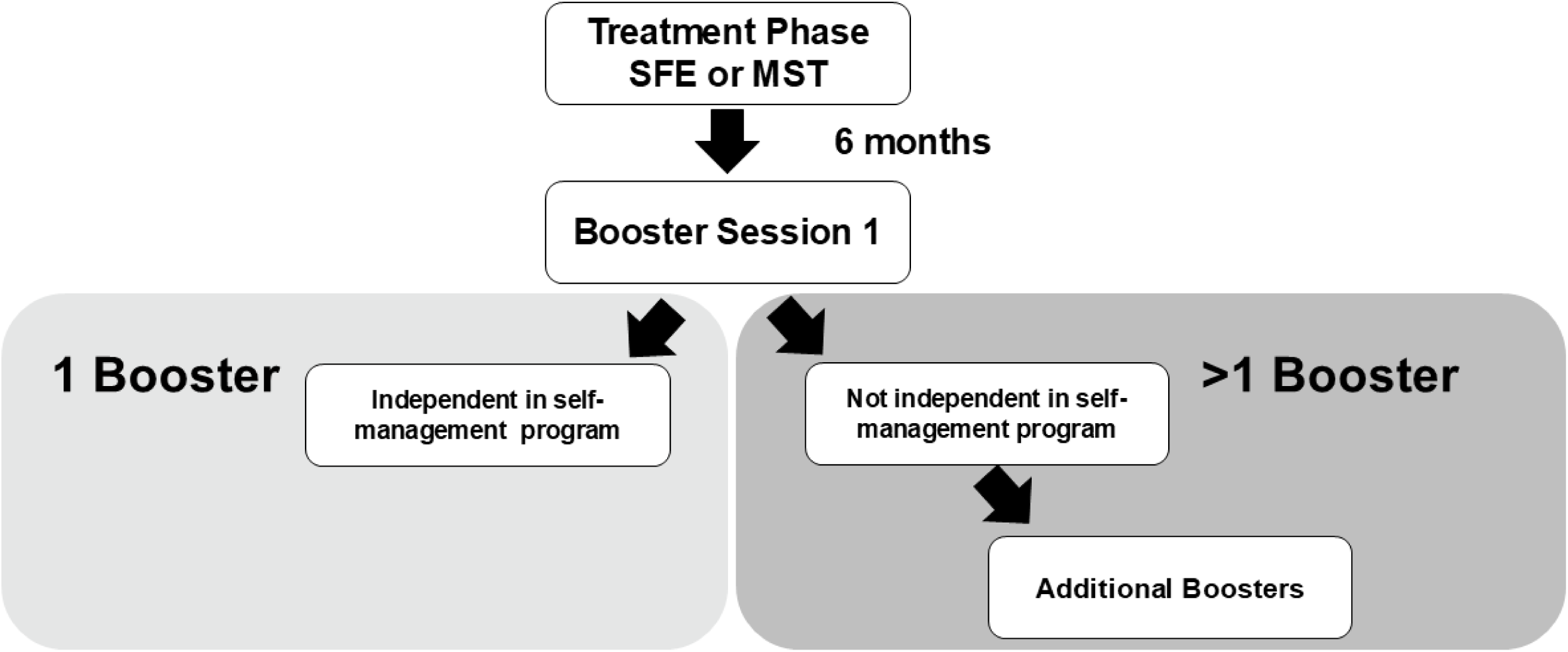
Flow chart illustrating the prescription of booster sessions at booster session 1. Participants who were independent with their self-management program at the first booster were prescribed 1 booster, and participants who were not independent were prescribed >1 booster.

## METHODS

### Design

This is a secondary analysis of a single-blind, RCT where participants with chronic, non-specific LBP were randomized to MST, MST+B, SFE, or SFE+B.^20^ All testing and treatment occurred at Washington University School of Medicine. The study was approved by the Washington University School of Medicine Institutional Review Board (IRB#: 201205051) and all participants provided informed consent. Individuals with chronic LBP (n=154) were recruited from the Saint Louis, Missouri, USA metropolitan area from December 2013 to August 2016. The sample size was determined for the primary aims of the RCT by an a priori power analysis to detect a difference of 6 points on the modified Oswestry Disability Questionnaire (MODQ)^21^ with 80% power.^20^

### Participants

154 participants between 18–60 years old with chronic non-specific LBP were enrolled in the trial. Full inclusion and exclusion criteria are in **TABLE 1**. The sample for this secondary analysis included the 76 participants who were randomized to boosters.

**TABLE 1.** Inclusion and exclusion criteria for the trial.

|  |
| --- |
| **Inclusion Criteria**<br>• 18–60 years old<br>• Chronic low back pain (LBP) for $\geq 12$ months<br>• Current LBP and not in an acute flare-up<br>• Modified Oswestry Disability Questionnaire $\geq 20\%$<br>• $\geq 3$ LBP-limited functional activities<br>• Able to stand and walk without assistance<br>• Able to comprehend and read English and sign a consent form. |
| **Exclusion Criteria**<br>• BMI > 30<br>• Neurologic disease requiring hospitalization<br>• Active cancer<br>• LBP etiology other than the lumbar spine<br>• Pregnancy<br>• LBP-related worker's compensation, disability or litigation<br>• Fibromyalgia<br>• Marfan syndrome<br>• Graves' disease<br>• Inability to classify LBP<br>• Any specific cause of LBP<br>• History of spinal fracture or surgery<br>• Frank neurological loss<br>• Pain or paresthesia below the knee. |

### Intervention

In the initial treatment phase, participants received 6 weekly, 1-hour sessions of MST or SFE. MST was person-specific training using motor learning principles to change movement and alignment patterns during LBP-limited functional activities.^20,22,23^ SFE was exercises following ACSM guidelines to improve trunk muscle strength and trunk and lower limb flexibility in all planes. Full descriptions can be found in the primary outcome manuscript, Supplement 2.^20^ At each visit, participants were assigned a program of their prescribed treatment to perform between sessions. At each subsequent visit, a reliable performance-based measure (inter-rater reliability, performance: % = 91 and k_w_ = 0.72 [95% CI 0.47-0.97]) was used to assess if participants were independent in their program and could be progressed.^24^ Independence included the ability to (1) verbalize the key concept of the exercise/skill, (2) perform the exercise/skill correctly without cues for (3) the prescribed number of repetitions (**FIGURE 2**). At the final treatment phase visit, participants were prescribed a self-management program of exercises/skills in which they were independent and encouraged to continue the program regularly for long-term self-management of their LBP. Six months later, participants attended a laboratory visit where they were informed of their booster assignment (booster/no booster). The 6-month time point was chosen because that is when participants experienced a worsening of outcomes in a previous trial.^25^ This secondary analysis is focused only on those participants randomized to boosters to examine characteristics of those who may need >1 booster and the effects of additional boosters on function, pain, and adherence.

**FIGURE 2.**
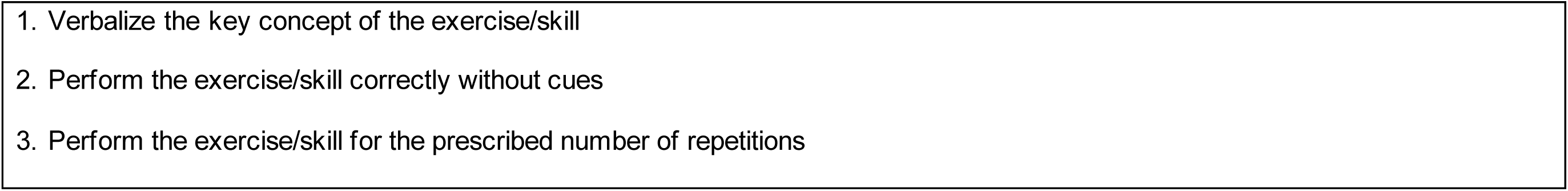
Criteria for independence in the self-management program. The participant is independent if all three criteria are met. SFE example for the exercise *side plank on elbow and foot*, 1. Participant verbalizes “the purpose of the exercise is to strengthen the low back and abdominals”, 2. Performs *side plank on elbow and foot* without cues, 3. For the prescribed number of repetitions, 3 sets of 10 repetitions. MST example for the skill *picking up groceries from a cart and placing them in the trunk of the car*, 1. Participant verbalizes “the key concept of the skill is to bend more in my hips and knees and less in my back as I pick up and place the groceries”, 2. Performs *picking up groceries from a cart and placing them in the trunk of the car* correctly, 3. For the prescribed number of repetitions, 2 sets of 10 repetitions.

At the first booster, each participant’s independence in their self-management program was assessed using the same performance-based measure used during the treatment phase (**FIGURE 2**). If the participant was independent, no additional boosters were prescribed. If the participant was not independent, then up to 2 additional sessions were prescribed weekly until the participant was independent or reached the 3-session maximum (**FIGURE 1**). We chose self-management program independence as the criterion for determining who required >1 booster because a primary aim of boosters is to help promote long-term behavior change and self-management.^8,26–28^ The 3-session maximum was set to minimize costs while providing enough treatment to boost self-management program independence. Treating therapists assessed the participants’ independence in their self-management program at the initial booster visit and were, therefore, not blinded to treatment assignment. However, the therapists that treated the MST group were not the same therapists as those that treated the SFE group.

### Measures

The primary outcome for the RCT was the MODQ, a validated measure of LBP-related functional limitation with higher scores indicating worse function.^21^ Secondary outcomes were the Numeric Pain Rating Scale (NRS; 0-10) for average and worst LBP in prior 7 days,^29^ adherence to the home program (participant-reported weekly adherence, 0-100%, during the treatment and booster phase and monthly adherence, 0-100%, during the follow-up phase),^25,30^ LBP flare-ups,^31,32^ SF-36 Physical and Mental Component Summary scores,^33–35^ absenteeism and presenteeism,^36,37^ LBP-related treatment, medication, and equipment use, fear-avoidance beliefs^38,39^ and satisfaction with care.^40^ All measures (except adherence to treatment and satisfaction with care) were collected at baseline. At each treatment session or booster session, MODQ, pain, and adherence to treatment (for visits 2-6 or boosters 2-3) were collected. After the initial treatment phase participants completed a subset of self-report surveys monthly for 12 months. All data were collected using REDCap.^41,42^ Outcome testers were blinded to treatment assignment. For this secondary analysis, additional measures included attendance at the first booster session and the number of boosters prescribed for those who attended the first booster.

### Statistical Analysis

#### Booster attendance and prescription

All statistical analyses were conducted in R v4.1.2.^43,44^ To analyze attendance at the first booster session (yes/no) and the number of boosters as a function of Group (MST versus SFE), we used a Chi-square test with Yates’ continuity correction, which reduces over estimation in small samples.^45^

#### Moderators of booster prescription

Next, we used logistic regression to model the probability of needing >1 booster for those participants who attended the first booster session (i.e., participants who did not attend any boosters were excluded from these analyses). Given our limited sample size, we started with a model testing the effect of booster prescription (>1 versus 1 session) regressed onto Group (MST versus SFE) as a base model. We then tested a series of individual models adding the effects of age, sex, duration of LBP, adherence, MODQ, and average pain as predictors controlling for group.

#### Exploring differences in outcomes in those who needed additional boosters

Finally, we were interested in how booster attendance might affect adherence, MODQ, and pain from the booster phase to the post-booster phase (the six months following the last booster) of the study. However, given the lack of statistical power and the small, uneven group sizes, we decided not to calculate inferential statistics for these changes. Instead, we present descriptive statistics for adherence, MODQ, and pain for participants who needed 1 booster versus >1 booster in each group (MST versus SFE) during the booster phase and the post-booster phase.

## RESULTS

### Participant characteristics at baseline at the beginning of the trial

The sample for these analyses included the 76 participants (average age: 40.1±11.2 years, 49 female) who were randomized to boosters. At the beginning of the trial the participants had an average MODQ of 30.7±9.5, average pain rating in the past week of 4.7±1.7, and LBP duration of 9.8±8.8 years (**TABLE 2**). Forty participants were randomized to MST+B and 36 were randomized to SFE+B. The characteristics of participants in the MST+B group and SFE+B group were not significantly different with the exception of sex and worst pain in the prior 7 days. There were significantly more females in the MST+B group (*p*=0.017) and the worst pain was significantly higher in the SFE+B group (*p*=0.044).

**TABLE 2.**
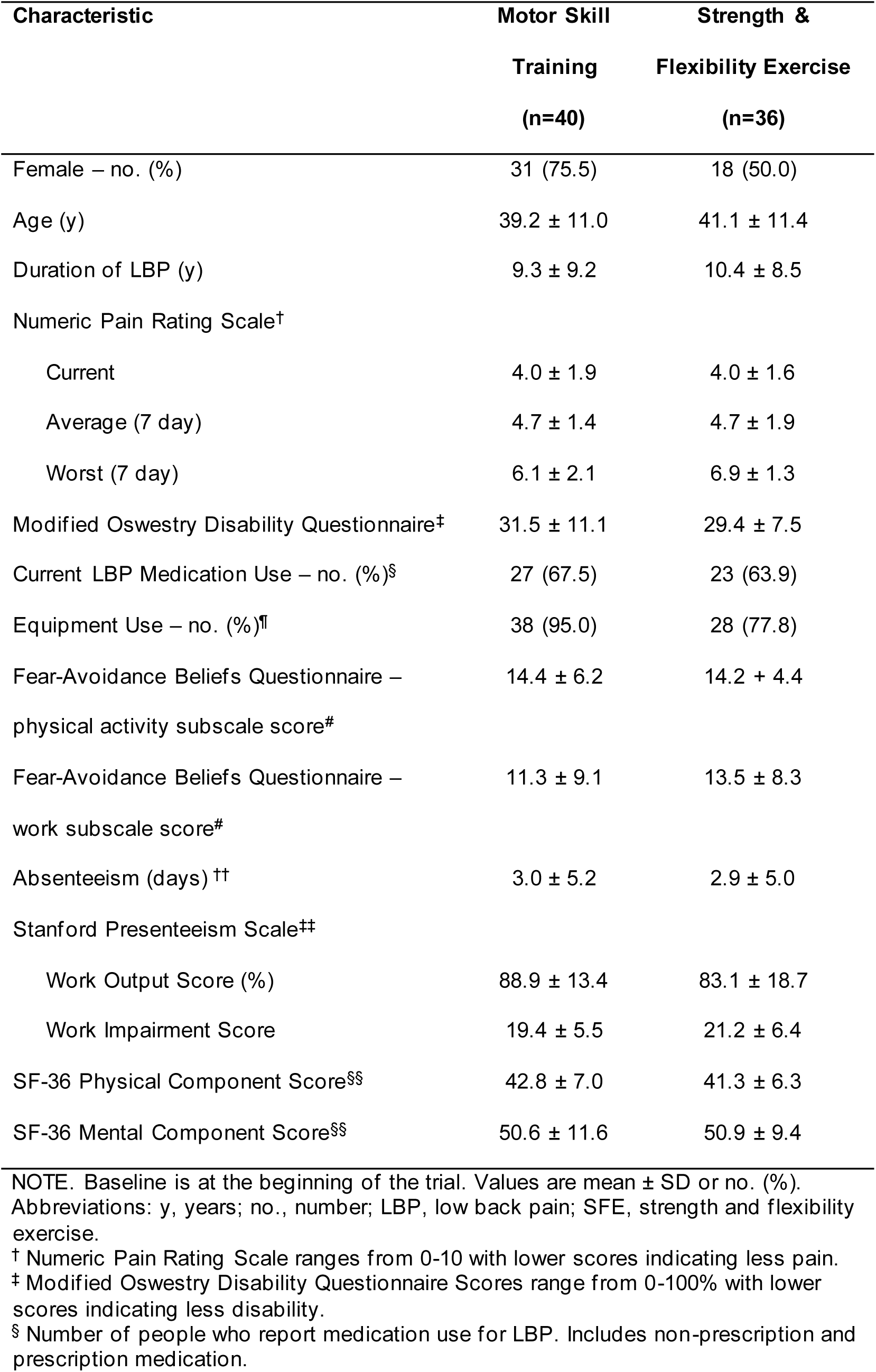
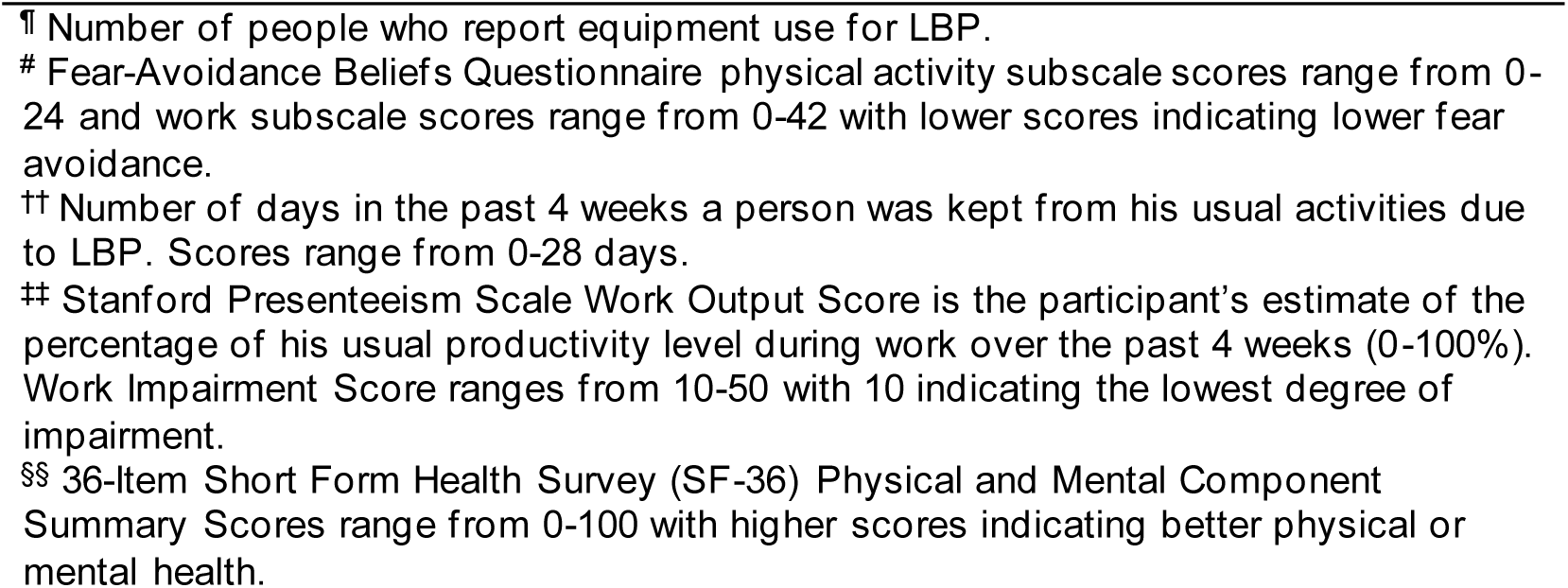
Baseline characteristics for individuals randomized to booster treatment.

| Characteristic | Motor Skill | Strength & |
| --- | --- | --- |
|  | Training<br>(n=40) | Flexibility Exercise<br>(n=36) |
| Female – no. (%) | 31 (75.5) | 18 (50.0) |
| Age (y) | 39.2 ± 11.0 | 41.1 ± 11.4 |
| Duration of LBP (y) | 9.3 ± 9.2 | 10.4 ± 8.5 |
| Numeric Pain Rating Scale<sup>†</sup> |  |  |
| Current | 4.0 ± 1.9 | 4.0 ± 1.6 |
| Average (7 day) | 4.7 ± 1.4 | 4.7 ± 1.9 |
| Worst (7 day) | 6.1 ± 2.1 | 6.9 ± 1.3 |
| Modified Oswestry Disability Questionnaire<sup>‡</sup> | 31.5 ± 11.1 | 29.4 ± 7.5 |
| Current LBP Medication Use – no. (%)<sup>§</sup> | 27 (67.5) | 23 (63.9) |
| Equipment Use – no. (%)<sup>¶</sup> | 38 (95.0) | 28 (77.8) |
| Fear-Avoidance Beliefs Questionnaire –<br>physical activity subscale score<sup>#</sup> | 14.4 ± 6.2 | 14.2 ± 4.4 |
| Fear-Avoidance Beliefs Questionnaire –<br>work subscale score<sup>#</sup> | 11.3 ± 9.1 | 13.5 ± 8.3 |
| Absenteeism (days) <sup>††</sup> | 3.0 ± 5.2 | 2.9 ± 5.0 |
| Stanford Presenteeism Scale<sup>‡‡</sup> |  |  |
| Work Output Score (%) | 88.9 ± 13.4 | 83.1 ± 18.7 |
| Work Impairment Score | 19.4 ± 5.5 | 21.2 ± 6.4 |
| SF-36 Physical Component Score<sup>§§</sup> | 42.8 ± 7.0 | 41.3 ± 6.3 |
| SF-36 Mental Component Score<sup>§§</sup> | 50.6 ± 11.6 | 50.9 ± 9.4 |

NOTE. Baseline is at the beginning of the trial. Values are mean ± SD or no. (%). Abbreviations: y, years; no., number; LBP, low back pain; SFE, strength and flexibility exercise.
<sup>†</sup> Numeric Pain Rating Scale ranges from 0-10 with lower scores indicating less pain.
<sup>‡</sup> Modified Oswestry Disability Questionnaire Scores range from 0-100% with lower scores indicating less disability.
<sup>§</sup> Number of people who report medication use for LBP. Includes non-prescription and prescription medication.
¶ Number of people who report equipment use for LBP.

### Booster attendance and prescription

As shown in **FIGURE 3**, of the 40 participants in the MST+B group, 33 returned for the first booster and 7 did not (5 discontinued trial participation prior to booster assignment, 1 refused to attend the 6-month lab visit but continued trial participation, and 1 refused boosters but continued trial participation). Of the 36 participants in the SFE+B group, 24 returned for the first booster and 12 did not (5 discontinued trial participation prior to booster assignment, 1 refused to attend the 6-month lab visit but continued trial participation, and 6 refused boosters but continued trial participation). The difference in returning for the first booster between groups was not statistically significant, *χ*^2^(1)=1.76, *p*=0.185. Among participants who returned for the initial booster, 27/33 (81.8%) MST+B required only 1 booster (i.e., were independent with their self-management program at the first session) compared to 7/24 (29.2%) SFE+B, a statistically significant difference χ^2^(1)=13.89, p<0.001. All of the participants in both groups were independent in self-management program performance in ≤ 3 booster visits except 1 SFE participant.

**FIGURE 3.**
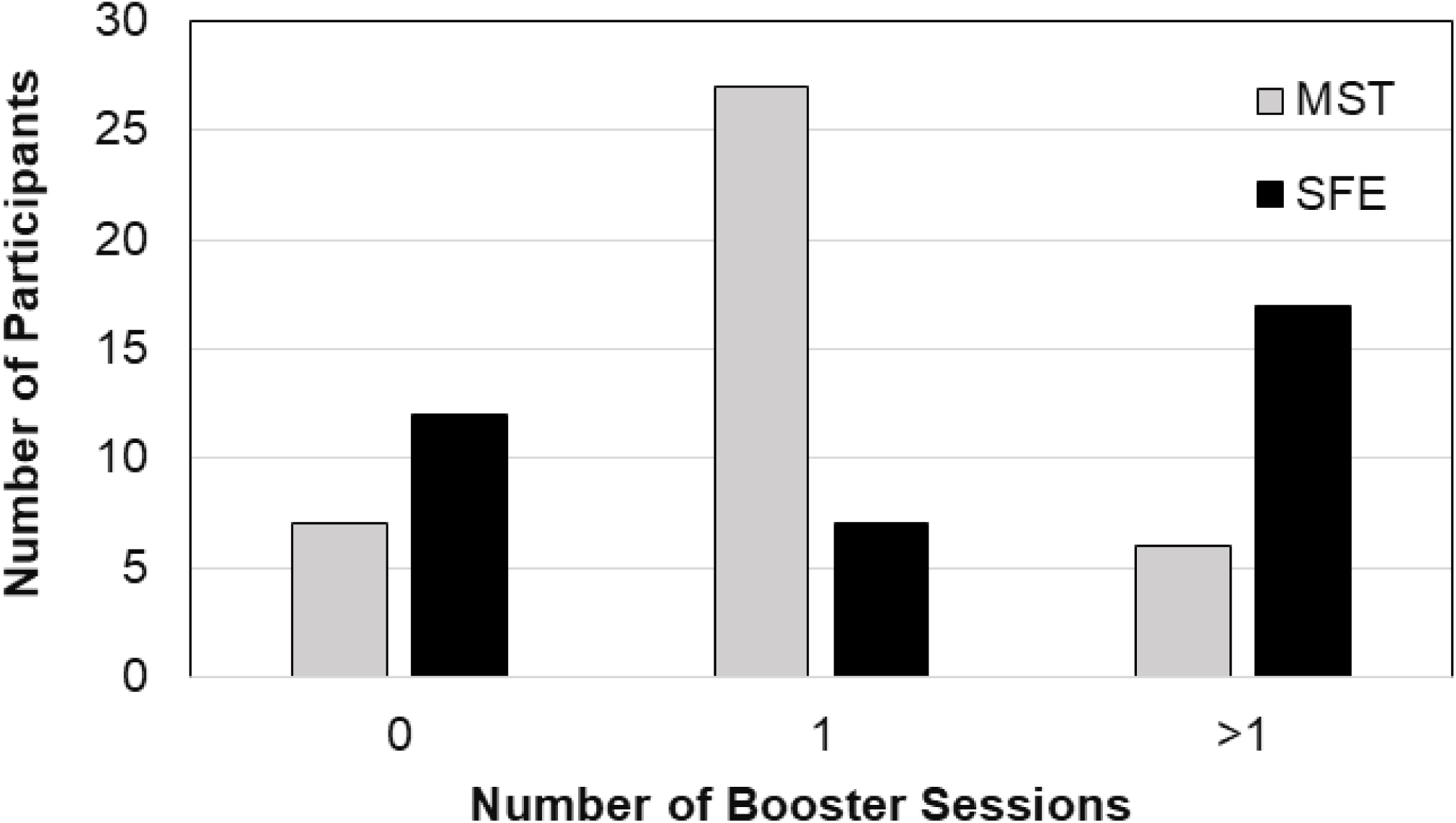
Number of booster sessions as a function of group. MST = motor skill training; SFE = strength and flexibility exercise. Participants who attended 0 booster sessions includes all participants randomized to boosters who did not attend the first booster session (randomized to boosters but discontinued trial participation prior to booster assignment, refused to attend the 6-month lab visit where they were informed of booster assignment, or refused boosters).

### Moderators of booster prescription

Logistic regression comparing who required >1 versus 1 booster again reflected that SFE+B participants were more likely to need to return for multiple sessions compared to MST+B participants, *β*=2.39, *p*<0.001, 95% CI, 1.14–3.64. Controlling for group, we added other variables in a series of individual models to see if they were related to requiring >1 booster. Neither age (*β*=0.04, *p*=0.173, 95% CI, −0.02-0.10), identifying as male (*β*=0.96, *p*=0.174, 95% CI, −0.42–2.34), the duration of LBP (*β*=0.03, *p*=0.450, 95%CI, −0.04–0.09), nor adherence to the self-management program prior to the booster (*β*=−0.01, *p*=0.386, 95%CI, −0.04–0.01) were significantly related to needing >1 booster. For this analysis, we were also able to look at how MODQ and pain at the first booster related to needing >1 booster. However, neither MODQ (*β*=0.02, *p*=0.490, 95%CI, −0.04–0.09) nor pain (*β*=0.21, *p*=0.388, 95%CI, −0.26–0.67) were significantly related to needing >1 booster.

### Exploring differences in outcomes in those who needed additional boosters

Descriptive statistics for adherence, MODQ, and pain for the booster phase and the post-booster phase (6 months following the booster) for those who required 1 and >1 booster are presented in **TABLE 3**.

**TABLE 3.** Descriptive statistics for adherence, modified Oswestry Disability Questionnaire (MODQ), and pain across the difference phases of the experiment.

| Group | Measurement |  |  |  |  |  |
| --- | --- | --- | --- | --- | --- | --- |
|  | Adherence<sup>†</sup> |  | MODQ<sup>‡</sup> |  | Average Pain (NRS)<sup>§</sup> |  |
|  | Booster | Post-Booster | Booster | Post-Booster | Booster | Post-Booster |
| *MST* |  |  |  |  |  |  |
| Required 1 Booster (n=27) | 73.1 (22.5) | 77.0 (17.6) | 9.4 (9.0) | 7.87 (7.4) | 1.19 (1.33) | 1.28 (1.14) |
| Required >1 Booster (n=6) | 75.2 (14.1) | 84.2 (9.8) | 14.0 (8.7) | 17.4 (17.9) | 1.67 (1.03) | 2.07 (1.62) |
| *SFE* |  |  |  |  |  |  |
| Required 1 Booster (n=7) | 46.9 (34.3) | 57.2 (32.1) | 20 (8.87) | 19.8 (9.0) | 2.14 (1.21) | 2.04 (1.20) |
| Required >1 Booster (n=17) | 31.4 (23.2) | 50.0 (17.8) | 19.5 (10.5) | 20.0 (13.2) | 2.41 (1.58) | 2.30 (1.63) |

Note that all values are presented as Mean (SD). For adherence, the “Booster” phase refers to adherence in the month immediately before the first booster session. For MODQ and average pain symptoms the “Booster” phase averages across all boosters attended, and then averages across participants. In the Post-Booster phase (the six months following the last booster), we averaged across all available data points within a subject, and then averaged across participants to get a group level average over time. MST = motor skill training; SFE = strength and flexibility exercise. <sup>†</sup>Adherence scores range from 0-100% with higher scores indicating better adherence. <sup>‡</sup>MODQ ranges from 0-100% with lower scores indicating less disability. <sup>§</sup>Numeric Pain Rating Scale (NRS) ranges from 0-10 with lower scores indicating less pain.

## Discussion

Our first purpose was to compare booster attendance and prescription in the MST and SFE groups. There was not a statistically significant difference in attending the first booster by group. However, there was a difference between groups in their independence at that session and thus in the number of boosters prescribed. The majority of MST+B participants were independent in their self-management program at the first booster session six months following the initial treatment phase (required 1 booster) whereas the majority of SFE+B participants required additional sessions (>1 booster) to attain independence. Current clinical practice guidelines emphasize the importance of self-management in chronic low back pain ^46^ — Our findings suggest that MST may result in better performance of a self-management program 6 months following treatment than SFE. This difference may be because MST was designed specifically to facilitate learning.

Our second purpose was to determine if there are variables that predict who will need >1 booster. When controlling for group, none of the participant-specific variables of age, sex, or duration of LBP were significantly related to needing >1 booster. Additionally, pain, function, and adherence were not significantly related to needing >1 booster. Since we did not find a relationship between needing >1 booster and LBP-related outcomes, one could question our criterion for determining who needs additional boosters. We used independence in the prescribed program as the criterion because the program was the primary method for LBP self-management. We did not use a change in pain or function to prescribe additional boosters, because the goal of the boosters was to help maintain self-management over the long-term rather than to wait for a worsening in pain or function. Our rationale for using self-management program independence is consistent with previous studies that describe the purpose of boosters as promoting long-term independence and behavior change.^13,14,16–19^ Different from our study, in previous studies of boosters for LBP the same number of sessions was prescribed for all participants.^11–19^ It is possible that a criterion other than self-management program independence would better predict who would benefit from >1 booster, for example, adherence, pain, or function – this could be explored in future studies because, to our knowledge, no other study has used a criterion to determine booster prescription.

We were not able to statistically test differences in outcomes for participants who needed >1 booster versus 1 booster. Thus, the values presented are a description of the current sample, rather than generalizations to the larger population. Qualitatively, in the SFE group, people who needed >1 booster look similar to people who needed 1 booster in terms of pain and function both during the booster and post-booster phase. It appears that for those who were not independent, additional treatment to “boost” their performance did not improve their pain or function. In contrast, in the MST group, people who needed only 1 booster qualitatively appear to be doing better than those who needed >1 booster, and better than people in the SFE group who attended any number of boosters. We previously reported that compared to no boosters, there was no effect of boosters on pain or function when including everyone who was randomized to boosters.^20^ Here we explore those that needed additional treatment per our criterion and there do not appear to be changes in pain or function in the 6 months following the booster. Therefore, it does not appear that boosters of the same treatment participants received during the initial intervention are beneficial, even when provided to people that are no longer independent in their prescribed method of self-management as in our study. It is possible that people need something different 6 months following treatment. Trials with innovative designs that provide additional treatment to non -responders during the initial treatment phase e.g., Sequential Multiple-Assignment Randomized Trials, are currently being explored in LBP and a similar approach could be explored for boosters.^47,48^

In addition to considering who may need additional boosters and what type of treatment should be prescribed, when to provide boosters is also a consideration. We chose 6 months following the initial intervention because that is when participants in a previous trial declined in function.^25^ However, participants in the current trial did not experience the same decline in function from 6-12 months.^20^ Perhaps boosters would have a greater effect if the timing of the boosters was individualized, since the changes in function observed in both trials were group averages rather than individual trajectories. This is particularly important to explore further since most previous studies of boosters in LBP provide no specific rationale for the timing of session s.^11,12,14–17,19^

### Limitations

We had a limited sample size to explore booster attendance and the lack of a significant difference in attending the first booster should not be interpreted robustly. For the moderators of booster prescription, the study was not powered to detect associations between additional boosters and prognostic variables, and these analyses are exploratory. Additionally, there were uneven and small numbers of participants in the groups who required 1 vs >1 boosters. Thus, we were unable to statistically explore differences between the groups. Since we specifically assessed MST and SFE, we cannot generalize our findings to other treatments for chronic LBP. However, SFE is one of the most recommended and commonly prescribed treatments for chronic LBP.^49^ Additionally, our findings may not be generalizable to individuals with LBP who have BMI >30, specific LBP conditions, or those receiving LBP-related worker’s compensation.

## Conclusions

There was not a statistically significant difference in returning for the first booster between the MST and SFE groups, but MST participants were more likely to be independent in their self-management program 6 months after treatment and thus needed fewer subsequent boosters than SFE participants. None of the additional factors we explored were significantly associated with self -management program independence. If long-term independent performance of a self-management program is a goal of LBP rehabilitation, then our findings suggest that clinicians may consider MST over SFE. While we were not able to statistically test differences in outcomes, qualitatively, attending additional boosters did not appear to change pain or function in the current sample. Our hypothesis-generating study suggests that future studies of boosters should examine the criteria for determining who needs additional treatment, the timing of sessions, and what type of treatment should be provided.

## Data Availability

All data produced in the present study are available upon reasonable request to the authors.

## Acknowledgements

The authors thank Kristen Roles, MS for assistance with study design and the members of the Musculoskeletal Analysis Group for their assistance with recruitment and data collection, processing, and analysis. The authors wish to acknowledge the Siteman Cancer Center’s National Cancer Institute (NCI) Cancer Center Support Grant P30 CA091842, the Washington University Institute of Clinical and Translational Sciences Grant UL1 TR002345 from the National Center for Advancing Translational Scien ces (NCATS), and the I2DB and Becker Library REDCap Support teams for supporting the Washington University instance of REDCap (Research Electronic Data Capture). NCI and NCATS are part of the National Institutes of Health (NIH).

